# Behavioral Effects of the California Flavored Tobacco Ban on Adult Cigarette Smoking: A Difference-in-Differences Analysis Using U.S. National Data

**DOI:** 10.64898/2026.01.13.26344037

**Authors:** Evan A. Winiger, Pavel N. Lizhnyak, Derek A. Pope, Andrea R. Vansickel

## Abstract

In December 2022, California enacted a comprehensive ban on the sale of flavored tobacco products, including menthol cigarettes. Using data from the Behavioral Risk Factor Surveillance System, this study employed difference-in-differences (DiD) models to examine adult smoking prevalence in California before (January 2017–December 2022) and after the ban (January 2023–December 2024), compared to states without flavor bans. From 2017-2024, smoking prevalence declined steadily in both California (from 11.8% to 7.9%) and the comparator states (from 18.5% to 12.4%). Adjusting for sociodemographic and year-fixed effects, the DiD estimate among adults 21+ was not significant (aOR=1.09; 95% CI: 1.00-1.20; p=0.0522). However, DiD effects showed significantly lower cigarette decline for age 21-34 (aOR=1.26; 95% CI: 1.05-1.52; p=0.0146) and for Hispanic adults (aOR=1.19; 95% CI: 1.01-1.41; p=0.0433). These results suggest that the flavor ban in California did not significantly affect overall adult 21+ smoking prevalence compared to states without such policies but may have reduced the decline in cigarette smoking among adults aged 21-34 and Hispanic individuals.

## Introduction

In December 2022, the state of California implemented a statewide ban on the sale of flavored tobacco products, including menthol cigarettes. To date, only a handful of limited studies have examined the impact of the California flavor ban on smoking-related outcomes. One study found that among a sample of California youth aged 12–17, past 30-day cigarette use dropped from 3.7% in 2022 to 2.8% in 2023; however, among those who smoked, 57% reported smoking menthol cigarettes in 2023 (1). Another 2023 study reported that 72.9% of a combined sample of California youth and young adults aged 13–24 still found menthol cigarettes easy to purchase (2), while a 2024 report indicated that only 21% of college students aged 18–24 were aware of the ban (3). Additionally, an analysis of sales data from 2019 to 2024 showed California’s flavor ban resulted in a near-complete drop in legal menthol sales, an average quarterly decrease in per capita cigarette sales, and an increase in tobacco-flavored e-vapor sales compared to control states without flavor bans. However, these legal retail data do not include illicit sales, lack behavioral interpretations, and the presence of flavor bans in a quarter of the control states complicates interpretation of the findings (4). Overall, existing research remains limited, focusing primarily on younger populations and lacking robust behavioral models that utilize pre- and post-ban trends.

Massachusetts is the only other state to implement a similar comprehensive ban on flavored tobacco products, which took effect in 2020. Behavioral studies have revealed unintended consequences associated with the Massachusetts flavor ban. One study utilizing a difference-in-differences (DiD) analysis on data from 2019 to 2023 found that the Massachusetts flavor ban was associated with increased cigarette use among young adults aged 18-24 and adults aged 25+ compared to states without flavor bans (5). A separate analysis using a DiD approach with Massachusetts data from 2017 to 2023 found that the state’s flavor ban had no significant impact on smoking rates among adults aged 21 and older when compared to states without such bans (6).

To date, no study has comprehensively examined the behavioral impact of the California flavor ban on adult smoking. This report addresses this gap by analyzing repeated cross-sectional data from the Behavioral Risk Factor Surveillance System (BRFSS) between 2017 and 2024 among adults aged 21 and older. We employ DiD analyses to compare changes in smoking prevalence before and after the flavor ban in California with those in states that lack flavor bans. Additionally, subgroup analyses examine demographic differences, including sex, age, race and ethnicity, and educational level. This report is the first behavioral analysis of the impacts of the California flavor ban on smoking prevalence in adults 21+.

## Methods

### Study Sample

We analyzed BRFSS data from 2017 to 2024 among adult smokers aged 21+. The study was exempt from IRB approval and informed consent, as per the Common Rule.^1^

### Measures

The primary outcome was past 30-day smoking prevalence. Demographic variables included age, sex, race and ethnicity, education, and household income.

### Statistical Analysis

Difference-in-differences (DiD) models were used to compare smoking prevalence among adults aged 21 and older in California, both before (January 2017 – December 2022) and after (post-December 2022) the ban, with 33 comparator states that did not have flavor bans. States excluded due to state-level flavor bans, local flavor bans, or incomplete survey data were Colorado, Florida, Georgia, Illinois, Maine, Maryland, Massachusetts, Minnesota, Montana, New Jersey, New York, North Dakota, Ohio, Rhode Island, Utah, and Wyoming (7, 8)^2^. Models adjusted for sociodemographic characteristics (age, sex, race and ethnicity, education, and household income) and included year-fixed effects. Student’s t-tests assessed differences in continuous demographic variables, and chi-square tests evaluated differences in categorical variables. We applied year-specific BRFSS survey weights to all analyses to produce population-representative estimates.

DiD models assessed (1) state-invariant changes in the odds of cigarette smoking from pre-to post-ban regardless of state flavor ban policy (time effect), (2) time-invariant differences in cigarette smoking between California and comparator states during the study period (treatment effect), and (3) the interaction between time and treatment effects, which reflects whether the pre—post ban change in cigarette smoking differed between California and comparator states, and estimates the difference-in-differences in smoking prevalence by comparing temporal pre-post ban changes in cigarette smoking in California relative to comparator states (interpreted as the effect of the flavor ban on cigarette smoking or the DiD estimate). Our DiD analyses report odds ratios (ORs) and include an unadjusted model, followed by models that adjust for sociodemographics, and then sociodemographics plus year-fixed effects.

A significant odds ratio < (>) 1 for the time effect indicates that the odds of smoking decreased (increased) from the pre-ban to the post-ban period, regardless of treatment. A significant odds ratio < (>) 1 for the treatment effect indicates relatively lower (higher) odds of smoking in California (or selected California subgroup) than comparator states when averaged over the full pre-post study period. A significant odds ratio < 1 for the DiD estimate indicates a greater reduction (or lower increase; or more favorable change) in pre-post cigarette smoking odds in California relative to comparator states, suggesting the flavor ban had the intended effect. A significant odds ratio > 1 for the DiD estimate indicates a greater increase (or smaller reduction; or less favorable change) in pre-post cigarette smoking odds in California relative to comparator states, suggesting the flavor ban had an unintended effect. A non-significant DiD estimate implies that there was no difference in pre-post-cigarette smoking odds in California relative to comparator states, indicating the flavor ban had no effect.

Several sensitivity analyses examined the robustness of the fully adjusted (sociodemographics, year-fixed effects) DiD model by: 1) including all states as comparators, 2) excluding states bordering CA (Oregon, Nevada, Arizona), 3) adjusting for border status (Mexico or Canada), 4) adjusting for e-cigarette tax status (yes/no) as of 2024, and 5) excluding 2020 data to indirectly account for the COVID-19 pandemic effects.

Finally, to further evaluate heterogeneity in the policy’s impact, domain-specific analyses were conducted by applying the fully adjusted DiD model within subgroups of age (21-34, 35-54, 55+), race/ethnicity (Non-Hispanic white, non-Hispanic black, Hispanic, non-Hispanic other), sex (male, female), and education (less than high school diploma, high school diploma, some college, at least a college degree).

## Results

**Table 1** summarizes demographic and other characteristics of participants from California and comparator states using combined BRFSS data from 2017-2024. The sample included 72,971 adults aged 21+ from California and 1,938,095 adults from comparator states. Cigarette smoking prevalence was significantly lower in California than in comparator states (9.9% vs. 15.8%). Relative to comparator states, California had a significantly higher proportion of Hispanic and NH-Other race/ethnicity individuals, a lower proportion of White individuals, more adults aged 21-34 and 35-54, fewer adults aged 55 and older, an overall younger mean population, and a higher proportion of individuals with household income ≥ $75,000.

**Table 1.** Cigarette Smoking and Sociodemographic Characteristics Among Adults 21+ in California and Comparator States in the 2017-2024 Behavioral Risk Factors Survey.

| Characteristic | California<br>(n=72,971) |  | Comparator States<br>(n= 1,938,095) |  | Mean<br>Diff | *p*-value |
| --- | --- | --- | --- | --- | --- | --- |
|  | % | 95% CI | % | 95% CI |  |  |
| **Cigarette Smoking** | 9.9 | (9.6, 10.2) | 15.8 | (15.7, 15.9) | 5.9 | **<0.0001** |
| **Sex** |  |  |  |  |  |  |
| Female | 50.9 | (50.4, 51.5) | 51.4 | (51.3, 51.6) | 0.5 | 0.0790 |
| Male | 49.1 | (48.5, 49.6) | 48.6 | (48.4, 48.7) | -0.5 |  |
| **Race and ethnicity** |  |  |  |  |  |  |
| NH-Black | 5.7 | (5.5, 6.0) | 12.1 | (12.0, 12.2) | 6.4 | **<0.0001** |
| Hispanic | 36.3 | (35.8, 36.8) | 13.8 | (13.6, 13.9) | -22.5 |  |
| NH-White | 38.1 | (37.6, 38.6) | 66.3 | (66.2, 66.5) | 28.2 |  |
| NH-Other | 19.9 | (19.4, 20.4) | 7.8 | (7.7, 7.9) | -12.1 |  |
| **Educational Level** |  |  |  |  |  |  |
| No HS diploma | 16.5 | (16.1, 16.9) | 11.9 | (11.8, 12.0) | -4.6 | **<0.0001** |
| HS diploma | 20.1 | (19.7, 20.6) | 27.6 | (27.5, 27.8) | 7.5 |  |
| Some college | 30.4 | (29.9, 30.9) | 31.1 | (31.0, 31.3) | 0.7 |  |
| ≥ College degree | 32.5 | (32.0, 32.9) | 28.9 | (28.7, 29.0) | -3.6 |  |
| Missing data | 0.5 | (0.4, 0.6) | 0.5 | (0.5, 0.5) | 0.0 |  |
| **Age** |  |  |  |  |  |  |
| 21-34 | 26.7 | (26.3, 27.2) | 25.3 | (25.2, 25.4) | -1.4 | **<0.0001** |
| 35-54 | 35.7 | (35.2, 36.2) | 33.9 | (33.8, 34.1) | -1.8 |  |
| 55+ | 37.6 | (37.0, 38.1) | 40.8 | (40.6, 40.9) | 3.2 |  |
| Mean Age | 48.5 | (48.3, 48.7) | 49.6 | (49.5, 49.6) | 1.1 | **<0.0001** |
| **Household income** |  |  |  |  |  |  |
| <\$10,000 | 5.4 | (5.2, 5.6) | 3.2 | (3.2, 3.3) | -2.2 | **<0.0001** |
| \$10,000 – 14,999 | 4.1 | (3.9, 4.3) | 3.1 | (3.0, 3.1) | -1.0 | |
| \$15,000 – 19,999 | 4.2 | (4.0, 4.4) | 4.6 | (4.5, 4.7) | 0.4 | |
| \$20,000 – 24,999 | 5.1 | (4.9, 5.3) | 6.1 | (6.0, 6.1) | 0.9 | |
| \$25,000 – 34,999 | 8.3 | (8.0, 8.6) | 9.1 | (9.0, 9.1) | 0.8 | |
| \$35,000 – 49,999 | 8.9 | (8.6, 9.2) | 11.1 | (11.0, 11.2) | 2.2 | |
| \$50,000 – 74,999 | 10.4 | (10.1, 10.7) | 13.1 | (13.0, 13.2) | 2.7 | |
| ≥75,000 | 35.7 | (35.2, 36.2) | 30.6 | (30.5, 30.8) | -5.0 |  |
| Missing data | 18.0 | (17.6, 18.5) | 19.2 | (19.0, 19.3) | 1.1 |  |

NH = Non-Hispanic; HS = High School
Boldface indicates statistical significance ( $p < .05$ )

Figure 1. illustrates the prevalence of cigarette smoking in California and comparator states from 2017 to 2024. Smoking prevalence declined from 18.5% to 12.4% in comparator states and from 11.8% to 7.9% in California. After the California flavor ban in December 2022, the decline slowed: prevalence decreased from an average of 10.4% pre-ban to 8.3% post-ban in California, compared to 16.7% pre-ban to 12.8% post-ban in comparator states, corresponding to a decrease of 1.8 percentage points less in California vs. comparator states. If all menthol smokers in California had quit following the ban (an estimated 30% (9)), the expected smoking prevalence in 2024 would have been approximately 5.5%.

**Figure 1.**
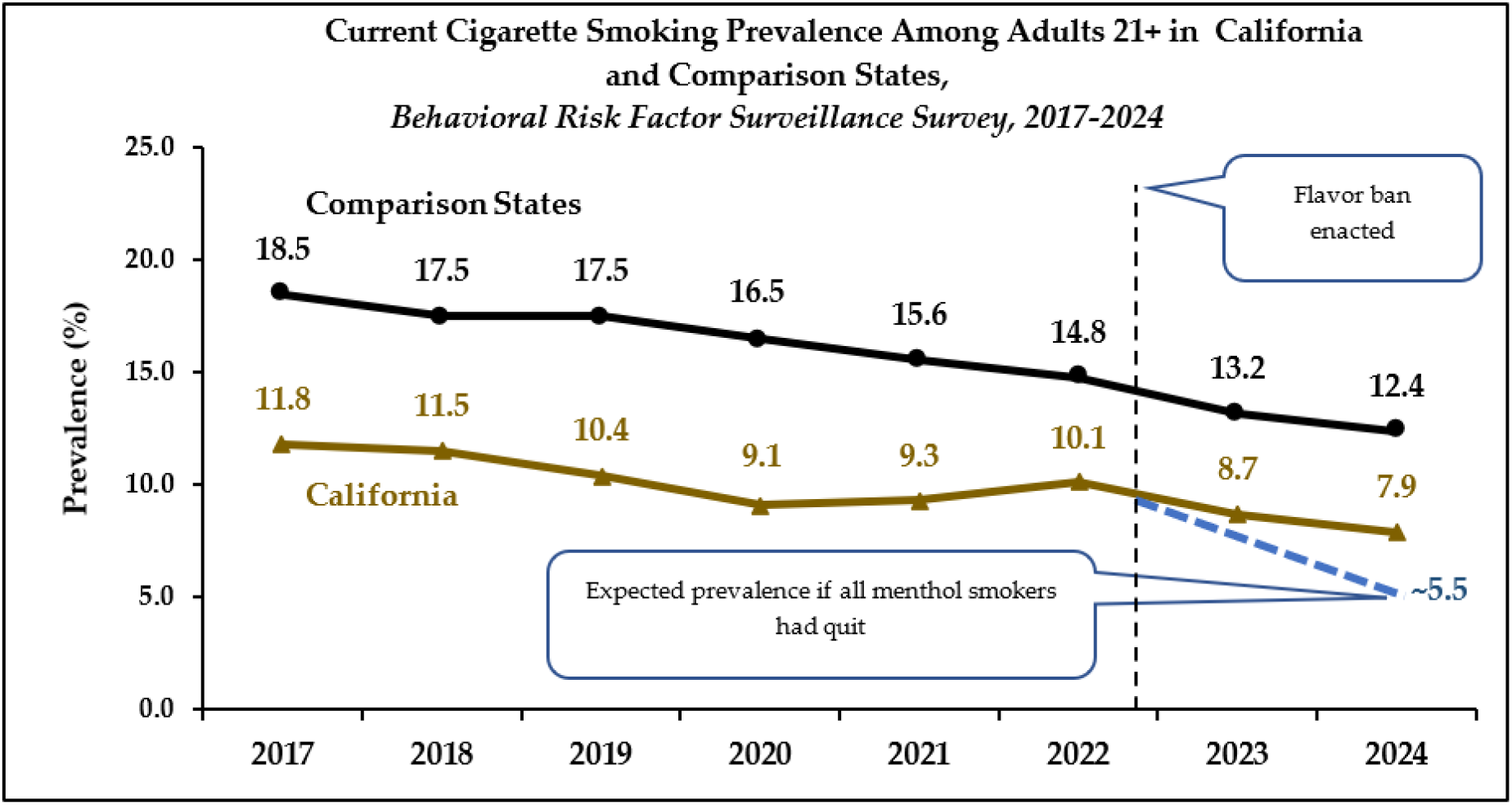
Prevalence of Cigarette Smoking in California and Comparator States from 2017-2024.

The parallel trends assumption was evaluated using multiple approaches, including visual inspection, unadjusted and adjusted parallel trends tests, and unadjusted and adjusted slope tests (with 2020 as the midpoint). The unadjusted parallel trends test was non-significant (p = 0.0511), whereas the adjusted test was significant (p = 0.0322), indicating small but statistically significant differences that reflect minor year-to-year variation. However, both the unadjusted and adjusted slope tests using 2020 as the midpoint showed no evidence of differential linear trends (*ps* > 0.5529). Taken together with visual inspection, these results suggest that pre-ban trends generally satisfy the DiD assumption (see also the sensitivity analyses below, excluding 2020; **Table 3**).

**Table 2.** Difference-in-Differences (DiD) Estimates from Unadjusted and Adjusted Models (Odds Ratios and 95% Confidence Intervals).

| Model | Change Over Time<br>(ref. pre-ban) |  | Changes Between Groups<br>(ref. Comparator States) |  | Comparison of Changes<br>Between Groups Over Time<br>(DID Estimate) |  |
| --- | --- | --- | --- | --- | --- | --- |
|  | aOR<br>(95% CI) | p -value | aOR<br>(95% CI) | p -value | aOR<br>(95% CI) | p -value |
| Unadjusted | 0.73 (0.72, 0.75) | <0.0001 | 0.58 (0.55, 0.60) | <0.0001 | 1.09 (1.00, 1.19) | 0.0566 |
| Adjusted for sociodemographics | 0.81 (0.80, 0.83) | <0.0001 | 0.68 (0.64, 0.71) | <0.0001 | 1.10 (1.01, 1.21) | **0.0354** |
| Adjusted for sociodemographics, year fixed effects | 1.00 (0.89, 1.12) | 0.9705 | 0.67 (0.64, 0.71) | <0.0001 | 1.09 (1.00, 1.20) | 0.0522 |

Boldface indicates statistical significance (p < .05)

**Table 3.**
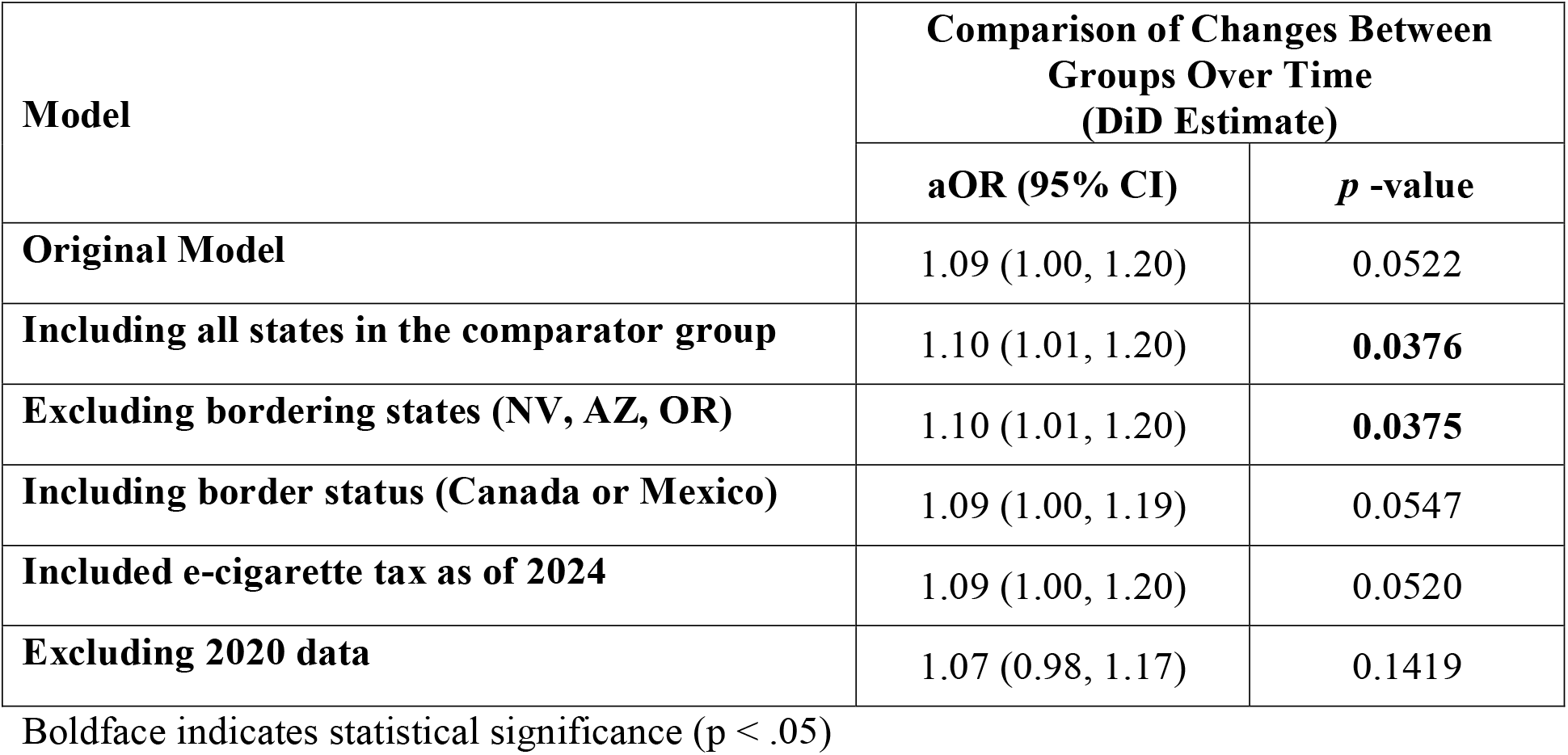
Sensitivity Analyses for Adjusted (sociodemographics, year fixed effects) Difference-in-Differences (DiD) Estimates (Odds Ratios and 95% Confidence Intervals).

**Table 2** presents results from three DiD models, including state-invariant changes in the odds of cigarette smoking over time (time effect), time-invariant differences in cigarette smoking between California and comparator (treatment effect), and the interaction between time and treatment effects (the DiD estimate) which provides the impact of the flavor ban on cigarette smoking when accounting for time and treatment effects. We first report the ORs with 95% confidence intervals (CIs) from the unadjusted model, followed by adjusted ORs (aORs) and 95% CIs from models also including sociodemographics, and then sociodemographics plus year-fixed effects. In the unadjusted model, time (OR=0.73; 95% CI: 0.72-0.75; p<0.0001) and treatment (OR=0.58; 95% CI: 0.55-0.60; p<0.0001) were significant and < 1, indicating that the odds of smoking decreased over time regardless of state policy and the odds of smoking were overall lower in California than comparator states from 2017-2024, respectively. The DiD estimate did not reach significance (OR=1.09; 95% CI: 1.00-1.19; p=0.0566). After adjusting for sociodemographics, time (aOR=0.81; 95% CI: 0.80-0.83; p<0.0001), treatment (aOR=0.68; 95% CI: 0.64-0.71; p<0.0001), and the DiD estimate (aOR=1.10; 95%CI: 1.01-1.21; p=0.0354) were all significant. Because the DiD estimate was significantly > 1 and the time and treatment effects were significantly < 1, this implies that while pre-post cigarette smoking odds decreased significantly, regardless of state policy, and was significantly lower overall in California compared to comparator states, the reduction in odds of cigarette smoking from pre-to post-ban was significantly less in California compared to comparator states. Further adjustment for year-fixed effects rendered time (aOR=1.00; 95% CI: 0.89-1.12; p=0.9705) and the DiD estimate non-significant (aOR=1.09; 95% CI: 1.00-1.20; p=0.0522), however, treatment (aOR=0.67; 95% CI: 0.64-0.71; p<0.0001) remained significant. In the fully adjusted model, there was a significant time-invariant difference in overall smoking rates between California and comparator states (treatment effect), but there was no significant state policy-invariant change in cigarette smoking trends over time (time effect), nor did the pre-post change in odds of cigarette smoking differ in California relative to comparator states, indicating the flavor ban had no impact on cigarette smoking (the DiD effect).

We evaluated robustness of our findings on the DiD estimates through a series of sensitivity analyses. These analyses yielded aORs ranging from 1.07 to 1.10, with overlapping 95% CIs and p-values ranging from 0.0375 to 0.1419 (**Table 3**). Across all specifications, the estimated direction and magnitude of the DiD aORs remained stable, while nominal statistical significance was sensitive to model choices near the conventional 0.05 threshold.

All sociodemographic variables in the two adjusted DiD models were significant, suggesting the need to assess the policy impact within each sociodemographic subgroup. **Table 4** shows the results of domain-specific analyses using the fully adjusted DiD models (sociodemographics, year-fixed effects) across each level of age, race/ethnicity, sex, and education. The domain analyses revealed that across age and race/ethnicity subgroups, the DiD parameter estimates were significant and positive (aOR > 1.0) among only 21-34-year-olds (aOR=1.26; 95%CI: 1.05-1.52; p=0.0146) and those of Hispanic ethnicity (aOR=1.19; 95%CI: 1.01-1.41; p=0.0433). The significant and positive DiD aORs indicated a relative increase in the pre—post odds of cigarette smoking among these California subgroups, effectively reducing the pre-post decline in cigarette smoking compared to these groups in comparator states. For all subgroups across age, race/ethnicity, sex, and education, the domain-specific DiD models revealed no significant effect of time (all p > 0.077), consistent with the DiD model adjusted for sociodemographics plus fixed-year effects applied to the full adult population aged 21 and above. Similarly, aORs for treatment were significant (i.e., aOR < 1.0) for all subgroups across age, sex, and education, and for Hispanic, NH-White, and NH-Other race/ethnicity (all aORs=0.58-0.76; all 95%CIs: 0.54-0.83; all p-values < 0.0001), but was not significant for those of NH-Black race/ethnicity (aOR=0.95; 95%CI: 0.80-1.14; p=0.5937). All significant treatment aORs were < 1.0, indicating that cigarette smoking was overall lower for the subgroups mentioned above in California relative to comparator states.

**Table 4.** Difference-in-Differences (DiD) Estimates from Subgroup-specific Adjusted (sociodemographics, year fixed effects) Models (Odds Ratios and 95% Confidence Intervals)

| Model | Change Over Time<br>(*ref. pre-ban*) |  | Changes Between Groups<br>(*ref. Comparator States*) |  | Comparison of Changes<br>Between Groups Over<br>Time<br>(**DID Estimate**) |  |
| --- | --- | --- | --- | --- | --- | --- |
|  | aOR<br>(95% CI) | *p* -value | aOR<br>(95% CI) | *p* -value | aOR<br>(95% CI) | *p* -value |
| **Among Age Levels** |  |  |  |  |  |  |
| 21-34 | 0.85 (0.63, 1.14) | 0.2775 | 0.65 (0.60, 0.71) | **<0.0001** | 1.26 (1.05, 1.52) | **0.0146** |
| 35-54 | 1.04 (0.86, 1.26) | 0.6877 | 0.68 (0.63, 0.73) | **<0.0001** | 1.08 (0.93, 1.25) | 0.3052 |
| 55+ | 1.07 (0.92, 1.23) | 0.3947 | 0.67 (0.62, 0.73) | **<0.0001** | 1.03 (0.89, 1.18) | 0.7057 |
| **Among Race/ Ethnicity Levels** |  |  |  |  |  |  |
| NH-Black | 1.07 (0.83, 1.37) | 0.5958 | 0.95 (0.80, 1.14) | 0.5937 | 0.77 (0.56, 1.06) | 0.1054 |
| Hispanic | 1.35 (0.97, 1.89) | 0.0773 | 0.62 (0.57, 0.68) | **<0.0001** | 1.19 (1.01, 1.41) | **0.0433** |
| NH-White | 0.91 (0.81, 1.02) | 0.1059 | 0.73 (0.69, 0.78) | **<0.0001** | 1.05 (0.93, 1.19) | 0.4264 |
| NH-Other | 0.82 (0.57, 1.17) | 0.2687 | 0.65 (0.57, 0.74) | **<0.0001** | 0.95 (0.73, 1.22) | 0.6755 |
| **Among Sex Levels** |  |  |  |  |  |  |
| Male | 1.01 (0.87, 1.18) | 0.8998 | 0.75 (0.70, 0.79) | **<0.0001** | 1.07 (0.96, 1.20) | 0.2382 |
| Female | 0.99 (0.83, 1.18) | 0.9294 | 0.58 (0.54, 0.63) | **<0.0001** | 1.12 (0.96, 1.30) | 0.1386 |
| **Among Education Levels** |  |  |  |  |  |  |
| No HS diploma | 1.18 (0.82, 1.70) | 0.3726 | 0.68 (0.61, 0.77) | **<0.0001** | 1.11 (0.90, 1.37) | 0.3469 |
| HS Diploma | 0.93 (0.79, 1.09) | 0.3771 | 0.63 (0.58, 0.69) | **<0.0001** | 1.16 (0.98, 1.36) | 0.0798 |
| Some College | 0.95 (0.77, 1.18) | 0.6651 | 0.73 (0.67, 0.79) | **<0.0001** | 1.05 (0.90, 1.24) | 0.5085 |
| ≥College Degree | 1.04 (0.83, 1.30) | 0.7594 | 0.76 (0.70, 0.83) | **<0.0001** | 0.93 (0.78, 1.11) | 0.4281 |

NH = Non-Hispanic; HS = High School
Boldface indicates statistical significance ( $p < .05$ )

## Discussion

Results of this analysis indicate that the California comprehensive flavor tobacco ban had no significant effect on smoking prevalence among adults aged 21 and older compared to states without such bans. After adjusting for sociodemographic and year-fixed effects, the DiD estimate was not significant, suggesting no differences in the odds of smoking in California relative to comparator states following the flavor ban. These findings align with prior research on Massachusetts, which also reported no significant change in adult smoking rates following its flavored tobacco ban (6). It is worth noting that in the fully adjusted DiD model, the DiD estimate was marginally significant and > 1.0, indicating a reduction in the decline in adult smoking in California relative to comparator states following the implementation of the flavor ban.

Considering the significant DiD estimate in the subgroup analyses, the ban was associated with relatively smaller declines in smoking among California adults aged 21-34 and Hispanic individuals compared to these subgroups in states without bans. These patterns mirror Massachusetts findings, where younger adults aged 18-24 showed increased smoking after that flavor ban (5), and are consistent with evidence suggesting elevated smoking risk among Hispanic populations after flavor bans (10). Younger adults are most likely to initiate smoking (11, 12) and face long-term health risks and tobacco dependence, while Hispanic smokers are less likely to access smoking cessation treatments (13, 14). Additionally, minorities are more likely to obtain menthol cigarettes through illicit channels post-ban (15), increasing exposure to criminal activity and health disparities.

An unintended consequence of flavor bans may be reciprocal substitution, where limiting flavored e-vapor may lead to increased cigarette use. For example, a comparative study found that dual users of cigarettes and e-vapor in Massachusetts had significantly lower quitting smoking rates (16%) than those in states without bans (32%) (16). Multiple DiD studies have similarly reported relative increases in cigarette use in states with either comprehensive or e-vapor flavor bans (5, 10, 17, 18), suggesting these policies may reverse the long-term decline in cigarette smoking.

Other behavioral responses may include continued menthol cigarette smoking or switching to non-menthol cigarettes. Surveys and retail data from Massachusetts indicate that many smokers accessed menthol products from neighboring states or switched to non-menthol cigarettes (15, 19). A meta-analysis found that, following menthol bans, 50% of smokers switched to non-menthol cigarettes and 24% continued menthol use (20). In California, several tobacco companies introduced menthol workaround cigarettes that contained synthetic cooling agents after the ban (21, 22), which may have further undermined the effectiveness of policy.

Strengths of this study include the use of repeated cross-sectional nationally representative data gathered both before and after the implementation of the statewide flavor ban, which captures behavioral responses more comprehensively than sales data. This study has several limitations. First, BRFSS data are self-reported, which may introduce reporting bias. Second, the models may not fully account for other state-level tobacco policies, such as smoke-free laws or taxes on non-cigarette products. Third, menthol workaround cigarettes were not covered under the flavor ban, and their availability could have influenced smoking behaviors. Lastly, BRFFS lacks detailed measures on menthol use, menthol workarounds, non-menthol cigarette switching, and consistent e-vapor data, limiting the ability to assess additional behavioral responses to the ban.

This study provides the first behavioral analysis of the California flavored tobacco product ban on smoking in a representative adult 21+ population. Findings suggest that prohibiting flavored tobacco products, including menthol cigarettes, does not easily alter cigarette demand among adult smokers in California when compared to states without the ban, and may disproportionately impact younger and Hispanic populations negatively, potentially widening health disparities. Future policies should consider these unintended consequences and monitor behavioral responses to ensure equitable public health outcomes.

## Data Availability

All data produced are available online at https://www.cdc.gov/brfss/annual_data/annual_data.htm

## Footnotes

1 Part 46-protection of human subjects. Code of Federal Regulations. https://www.ecfr.gov/on/2018-07-19/title-45/subtitle-A/subchapter-A/part-46. Updated May 22, 2025. Accessed July 23, 2025.

2 While Oregon is listed by the Truth Initiative for having flavor bans in Multnomah and Washington County in 2024, <u>the Oregon Court of Appeals issued an order</u> that paused the implementation of the flavor ban in Multnomah County and the ban was not in effect by the end of 2024 and <u>the ruling in Washington County was moved to the Oregon Supreme Court</u> and did not take effect by the end of 2024. Thus, there were no active flavor bans in Oregon by the end of 2024, and the state was included as a comparator state.

